# Seroreversion to *Chlamydia trachomatis* Pgp3 antigen among children in a hyperendemic region of Amhara, Ethiopia

**DOI:** 10.1101/2023.02.04.23285360

**Authors:** Christine Tedijanto, Solomon Aragie, Sarah Gwyn, Dionna M. Wittberg, Taye Zeru, Zerihun Tadesse, Ambahun Chernet, Isabel J.B. Thompson, Scott D. Nash, Thomas M. Lietman, Diana L. Martin, Jeremy D. Keenan, Benjamin F. Arnold

## Abstract

Monitoring trachoma transmission with antibody data requires characterization of decay in IgG to *Chlamydia trachomatis* antigens. In a three-year longitudinal cohort in a high transmission setting, we estimated a median IgG half-life of 3 years and a seroreversion rate of 2.5 (95% CI: 1.6, 3.5) per 100 person-years.

## INTRODUCTION

Trachoma, caused by repeated conjunctival infection with *Chlamydia trachomatis*, is targeted for elimination as a public health problem by 2030. Control programs currently rely on clinical markers, including trachomatous inflammation—follicular (TF), an early indicator of inflammation, and trachomatis trichiasis, a sign of severe disease progression. However, clinical signs are prone to measurement error. PCR-detected chlamydia infections are an objective alternative, but as populations approach elimination infections become rare which poses a challenge for surveillance [1]. Antibody responses among children are being explored as an objective, sensitive marker of chlamydia exposure across transmission settings [2].

Population-level monitoring of trachoma using serology has focused on IgG responses to the Pgp3 antigen [2]. Studies have demonstrated marked increases in seroprevalence with age in trachoma endemic regions and limited antibody responses in populations near elimination [3–5]. A key summary statistic from the age-seroprevalence curve is the seroconversion rate, which is one measure of chlamydia force of infection [6]. In the absence of seroreversion, IgG seropositivity by age reflects the cumulative incidence of prior exposure to chlamydia infection in the population. Seroreversion is a change from seropositive to seronegative status, and, if common, then failing to account for seroreversion could bias estimates and the interpretation of standard analyses of age-seroprevalence curves. Few longitudinal studies have measured Pgp3 IgG decay over time, and all have been conducted in Tanzania’s Kongwa District. Seroreversion rate estimates range from 0 cases within six months after mass drug administration (MDA) of azithromycin [7] to 12.1% over 36 months (4 per 100 child-years) [8], 6.4% after one year (6.4 per 100 child-years) [9], to 4.0% after 6 months (8 per 100 child-years) [10].

Better characterization of seroreversion and the rate at which Pgp3 IgG levels decay among children across a range of transmission settings will aid the development and interpretation of seroepidemiologic models of trachoma transmission. Our goal was to estimate the decay rate of Pgp3 IgG levels and the seroreversion rate in a longitudinal cohort of children 1-9 years old in a hyperendemic region of Amhara, Ethiopia.

## METHODS

The WASH Upgrades for Health in Amhara (WUHA) cluster-randomized trial studied the effect of an integrated water, sanitation, and hygiene (WASH) intervention on ocular chlamydial infection (NCT02754583) [11]. The study enrolled 40 rural communities in the Wag Hemra Zone. Clinical disease, chlamydia infections, and serology were measured annually for three years in a longitudinal cohort of children aged 0–5 years at baseline. Approximately 30 randomly selected children per community were included, and infants under one year of age were newly enrolled in the longitudinal cohort at each annual visit. Once a child was enrolled in the longitudinal cohort, the study continued to test for infection and antibody responses even if they were 6–9 years.

Conjunctival swabs were collected from a random sample of 30 children per community at each visit, often including children in the longitudinal cohort. Swabs were tested for chlamydia DNA at the Amhara Public Health Institute in Bahir Dar, Ethiopia using the Abbott RealTime assay [11]. Dried blood spots were collected on TropBio filter paper and tested for IgG responses to Pgp3 in a multiplex bead assay on the Luminex platform using the same bead set for all samples. IgG was quantified using median fluorescence intensity minus background (MFI-bg) and responses >1113 were classified as seropositive using a receiver operator characteristic curve cutoff from reference samples [12]. Research was approved by the human subjects review board at the University of California, San Francisco. Each participant or guardian provided verbal consent before any study activity.

Before the trial, MDA was conducted annually for 7 years in the study area but had not sufficiently controlled trachoma [13]. MDA was suspended during the study period. Between baseline and 36 months, TF prevalence among 1-9 year-olds remained fairly constant from 63% to 57% [14] and ocular chlamydia infection among 0-5 year-olds increased from 11% to 32% with no difference between the control and intervention groups [11].

We estimated incidence rates non-parametrically and using Poisson regression with cluster-level random effects and an offset for person-years. Because this was an open cohort where children might have incomplete data across time points, we assessed serostatus over one-year intervals and only included one-year intervals with serology measurements at both time points. We assumed that changes in serostatus occurred halfway through the year. This analysis focused on the seroreversion rate (SRR), but we additionally estimated the seroconversion rate (SCR) using the same method. Measurements from children <1 year old were excluded from this analysis to avoid the influence of maternal antibody waning. We evaluated differences in SRR by age (1–5 years vs. 6–9 years), time period, and initial IgG level by adding covariates to the model. We estimated IgG half-life using an exponential decay model among children who began a one-year interval seropositive, ended the year with a negative or equivocal PCR test, and did not have an increase in IgG levels.

This analysis was conducted in R version 4.3.0 (2023-04-21). Data and replication files are available: https://osf.io/xquyd/.

## RESULTS

The analysis included 4,327 serology measurements from 1,511 unique children. We excluded 27 children who were determined to have mis-labeled blood specimens at one or more time points based on photography, anthropometry, and IgG levels. After filtering to one-year intervals with serology measurements at both ends, the analysis included 2,428 one-year intervals from 1,221 unique children (81%). Children who did not contribute were similar to those with partial (47%) or complete (33%) measurements over the 3-year period, but had more missing values for other trachoma indicators (**Supplementary Table 1**). Over the study period, seroprevalence among children in the longitudinal cohort increased from 30 to 51% as the cohort aged (**Supplementary Table 2**). The seroconversion rate (SCR) in the longitudinal cohort was 15.3 per 100 person-years (95% CI: 11.0, 20.8).

Seroreversion was rare: among 886 one-year intervals where children were seropositive at the beginning, 864 (98%) remained seropositive after one year. Among seropositive children, IgG levels remained high and were consistent with a durable response (**Figure 1A**), though there was some waning of IgG among children who were PCR negative or equivocal for chlamydia infection at the end of the interval (**Figure 1B**). There were 22 seroreversions during 875 seropositive person-years at risk, corresponding to a SRR of 2.5 per 100 person-years (95% CI: 1.6, 3.5). The estimated median IgG half-life was 3.0 years (IQR: 0.8, 9.6). Based on the slope of IgG decay among children who were seropositive at the beginning of the interval and PCR negative/equivocal at the end of the interval (**Figure 1B**), there appeared to be two decay rates, perhaps due to boosting between measurements or measurement error. Among the 20 children with >4 fold decrease in IgG levels (6 of whom seroreverted), median IgG half-life was 0.38 years (IQR: 0.27, 0.44). Seroreversion rates were lower: from months 24 to 36 of the study (SRR ratio compared to months 0 to 12: 0.26; 95% CI: 0.07, 0.97, **Figure 2A**) and among 6-9-year-olds (SRR ratio compared to 1–5-year-olds: 0.59; 95% CI: 0.17, 1.98, **Figure 2B**), though not statistically significant. Of 22 seroreversions, 14 were among children with IgG MFI-bg <10^3.5^ at the beginning of the interval with substantially lower seroreversion rates at higher IgG levels (**Figure 2C**).

**Figure 1.**
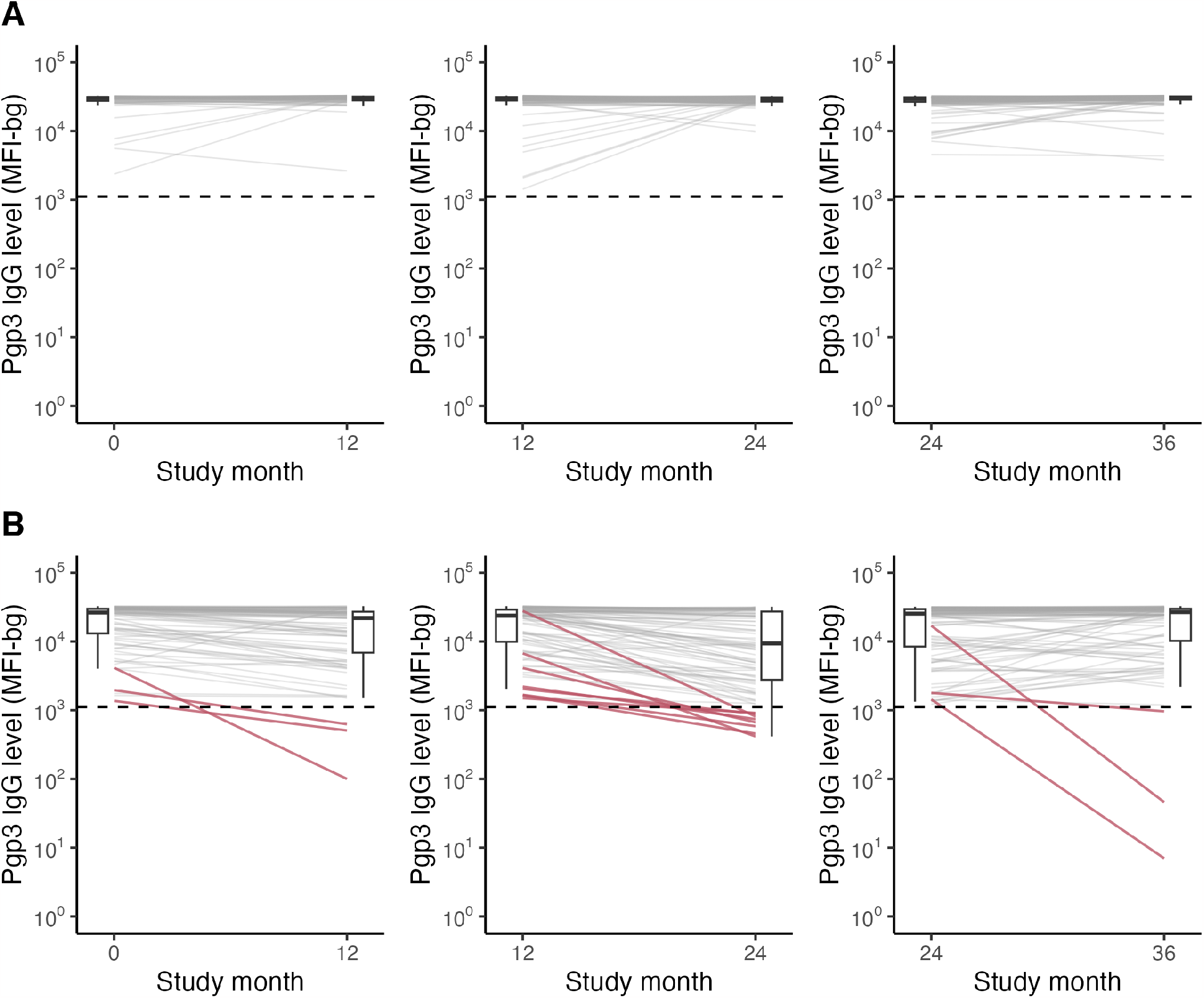
Antibody responses over time by C. trachomatis infection status. Pgp3 IgG responses among children who were seropositive at the beginning of each one-year interval and (A) PCR-positive compared to (B) PCR-negative or -equivocal for C. trachomatis infection at the end of the interval. Dashed horizontal line represents seropositivity cutoff of 1113 MFI-bg. Solid red lines in panel B identify children who seroreverted. Boxplots summarize median (thick horizontal line) and IQR (thin horizontal lines) at beginning and end of each interval. PCR results at the end of the interval were missing for 784 (32%) intervals due to random sampling for PCR testing in the cohort.

**Figure 2.**
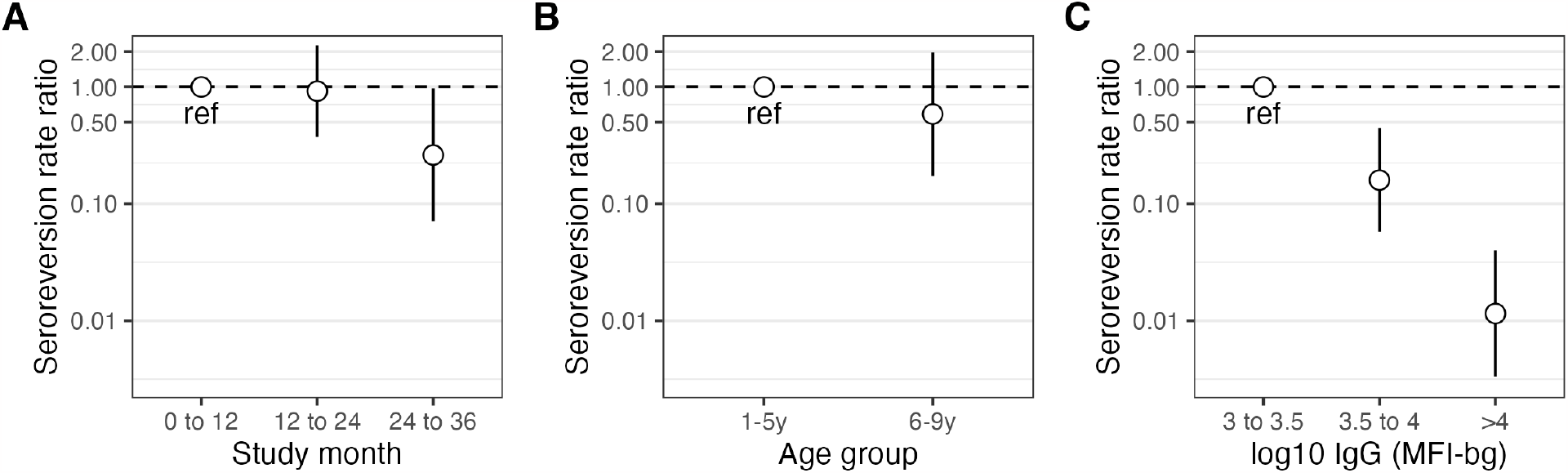
Seroreversion by subgroups. (A) Rate ratios and 95% CIs (on log scale) comparing seroreversion over time. (B) Rate ratios and 95% CIs (on log scale) comparing seroreversion by age group. (C) Rate ratios and 95% CIs (on log scale) comparing seroreversion by IgG level at the beginning of the one-year interval. Estimates underlying this figure are provided in Supplementary Table 3.

## DISCUSSION

In a hyperendemic region of Amhara, Ethiopia in the absence of MDA we found that seroreversion was rare and anti-chlamydia Pgp3 IgG antibody response was durable among children 1-9 years old, with a median half-life of 3 years. The IgG seroreversion rate was 2.5 per 100 person-years (95% CI: 1.6, 3.5) and seroreversion rates were lower at the end of the study amidst higher chlamydia transmission, among older children, and among children with higher initial IgG levels. To our knowledge, these are the first estimates of Pgp3 seroreversion rates in a high transmission setting. Strengths of the study include a well-characterized, three-year longitudinal cohort with paired measures that included clinical signs of trachoma, chlamydia infection measured with PCR, and IgG measured on a multiplex bead assay using a consistent bead set.

Our findings are similar to recent estimates of 6.4% seroreversion after one year in the absence of MDA [9] and 4% after 6 months in the presence of MDA in Kongwa, Tanzania [10], but may be slightly lower due to higher ongoing transmission (11% to 32% PCR prevalence) compared to Kongwa (4.9% to 6.3% PCR baseline prevalence). Higher SCR and lower SRR estimates in the present study likely reflect repeated infections that boost and maintain IgG levels above seropositivity cutoffs. Consistent with the results of our subgroup analyses, in Kongwa the SRR decreased with clinical disease activity and higher starting antibody levels.

This analysis had limitations. Children were followed longitudinally but some missed visits, leading to incomplete data. For this reason, we used an open cohort design and focused on changes in serostatus at one-year intervals to maximize follow-up time for analysis. If children who missed visits were systematically younger with higher seroprevalence, that could lead to a biased under-estimate of the SRR. Yet, children who went missing appeared similar based on age, seropositivity status, and other trachoma markers at enrollment to those with complete follow-up (**Supplementary Table 1**). A child could have seroconverted and reverted between annual visits. Although unlikely given the estimated 3-year IgG half-life, if transient changes in serostatus occurred between annual measurements, perhaps more likely amongst the youngest children [8], it would lead to an underestimate of seroconversion and seroreversion rates. Nevertheless, if trachoma control programs monitor serology with at most an annual frequency, the estimates presented in this study should be informative. Since the cohort aged over time, we cannot fully disentangle the effects of age and study month on the seroreversion rate. Finally, this study was conducted in a hyperendemic context with increasing transmission so findings may not generalize to lower transmission settings.

Trachoma programs are considering the use of serology to monitor transmission and recrudescence as populations approach elimination. In hyperendemic settings, serological measures of transmission serve as a baseline against which progress can be measured. In this longitudinal cohort, 2-3% of seropositive children experienced seroreversion of IgG responses to chlamydia antigen Pgp3 each year, demonstrating that seroreversion is unlikely to influence estimates of seroconversion rates among young children in hyperendemic settings.

## Data Availability

Data and replication files are available: https://osf.io/xquyd/

## ACKNOWLEDGEMENTS

The authors would like to thank Abbott for its donation of the m2000 Realtime molecular diagnostics system and consumables.

## SUPPLEMENTAL MATERIAL

**Supplementary Table 1.**
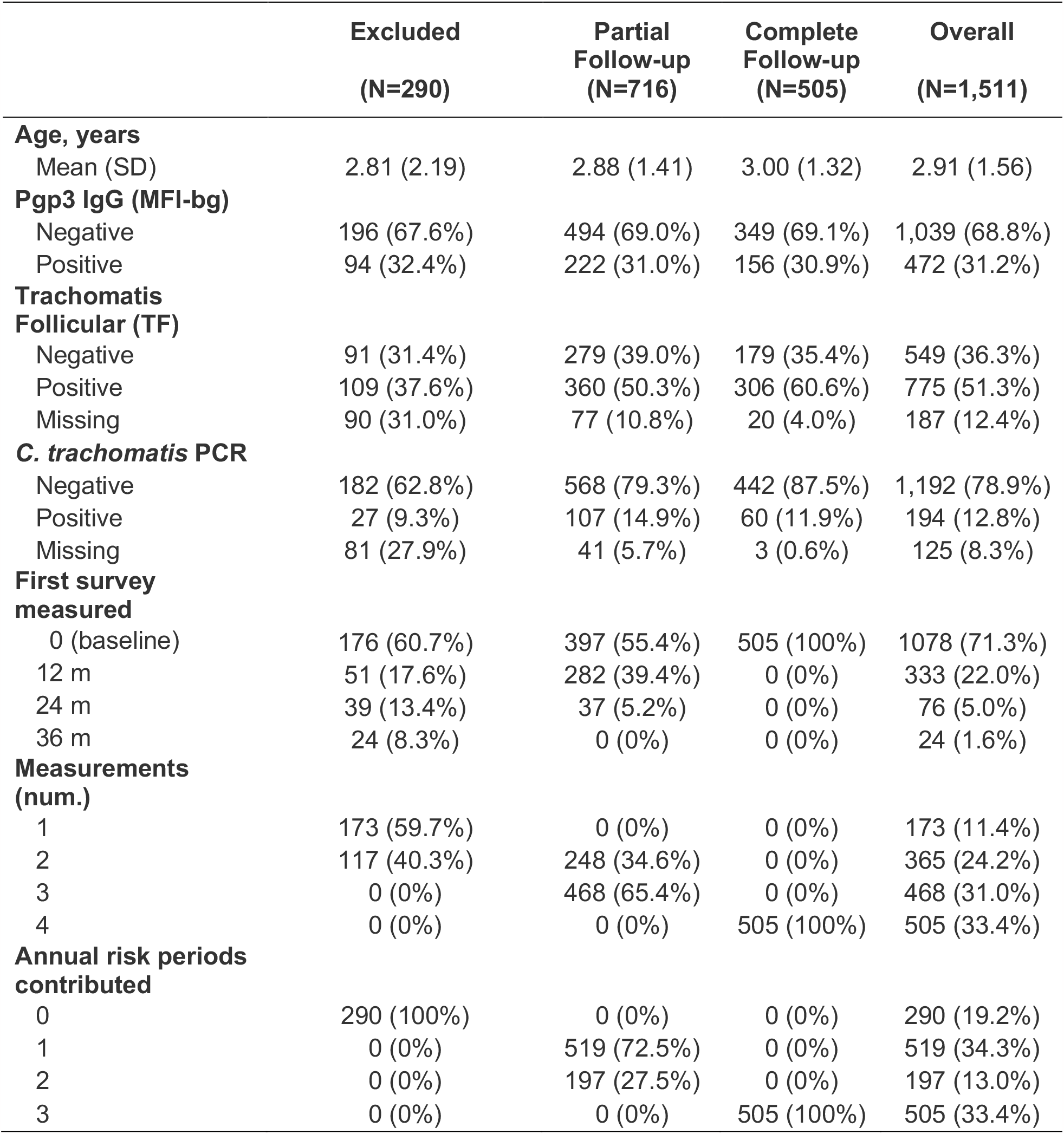
Summary of child characteristics in the longitudinal cohort at enrollment, number of measurements and annual periods contributed to the analysis by follow-up status. Children who did not contribute to the analysis (Excluded) did not have two adjacent serology measurements in annual surveys. Children who had complete follow-up were measured four times and contributed to three annual risk periods.

|  | Excluded<br>(N=290) | Partial<br>Follow-up<br>(N=716) | Complete<br>Follow-up<br>(N=505) | Overall<br>(N=1,511) |
| --- | --- | --- | --- | --- |
| **Age, years** |  |  |  |  |
| Mean (SD) | 2.81 (2.19) | 2.88 (1.41) | 3.00 (1.32) | 2.91 (1.56) |
| **Pgp3 IgG (MFI-bg)** |  |  |  |  |
| Negative | 196 (67.6%) | 494 (69.0%) | 349 (69.1%) | 1,039 (68.8%) |
| Positive | 94 (32.4%) | 222 (31.0%) | 156 (30.9%) | 472 (31.2%) |
| **Trachomatis**<br>**Follicular (TF)** |  |  |  |  |
| Negative | 91 (31.4%) | 279 (39.0%) | 179 (35.4%) | 549 (36.3%) |
| Positive | 109 (37.6%) | 360 (50.3%) | 306 (60.6%) | 775 (51.3%) |
| Missing | 90 (31.0%) | 77 (10.8%) | 20 (4.0%) | 187 (12.4%) |
| ***C. trachomatis* PCR** |  |  |  |  |
| Negative | 182 (62.8%) | 568 (79.3%) | 442 (87.5%) | 1,192 (78.9%) |
| Positive | 27 (9.3%) | 107 (14.9%) | 60 (11.9%) | 194 (12.8%) |
| Missing | 81 (27.9%) | 41 (5.7%) | 3 (0.6%) | 125 (8.3%) |
| **First survey**<br>**measured** |  |  |  |  |
| 0 (baseline) | 176 (60.7%) | 397 (55.4%) | 505 (100%) | 1078 (71.3%) |
| 12 m | 51 (17.6%) | 282 (39.4%) | 0 (0%) | 333 (22.0%) |
| 24 m | 39 (13.4%) | 37 (5.2%) | 0 (0%) | 76 (5.0%) |
| 36 m | 24 (8.3%) | 0 (0%) | 0 (0%) | 24 (1.6%) |
| **Measurements**<br>**(num.)** |  |  |  |  |
| 1 | 173 (59.7%) | 0 (0%) | 0 (0%) | 173 (11.4%) |
| 2 | 117 (40.3%) | 248 (34.6%) | 0 (0%) | 365 (24.2%) |
| 3 | 0 (0%) | 468 (65.4%) | 0 (0%) | 468 (31.0%) |
| 4 | 0 (0%) | 0 (0%) | 505 (100%) | 505 (33.4%) |
| **Annual risk periods**<br>**contributed** |  |  |  |  |
| 0 | 290 (100%) | 0 (0%) | 0 (0%) | 290 (19.2%) |
| 1 | 0 (0%) | 519 (72.5%) | 0 (0%) | 519 (34.3%) |
| 2 | 0 (0%) | 197 (27.5%) | 0 (0%) | 197 (13.0%) |
| 3 | 0 (0%) | 0 (0%) | 505 (100%) | 505 (33.4%) |

**Supplementary Table 2.** Sample size, age distribution, and seroprevalence in the longitudinal cohort, by month of the study.

| Study month | Total number of children with serology samples | Median age in months (IQR) | Seroprevalence (%) |
| --- | --- | --- | --- |
| 0 | 1,078 | 36 (24-48) | 31.4 |
| 12 | 1,155 | 48 (35-60) | 37.6 |
| 24 | 1,015 | 60 (47-72) | 42.2 |
| 36 | 1,079 | 72 (60-96) | 51.3 |

**Supplementary Table 3.** Seroconversion rate (SCR) and seroreversion rate (SRR) estimates overall and by subgroups. Children who were Pgp3 negative (–) at the beginning of a one-year period were at risk for seroconversion, and those who were Pgp3 positive (+) were at risk for seroreversion.

|  | Seroconversion Rate* |  |  |  | Seroreversion Rate* |  |  |  |
| --- | --- | --- | --- | --- | --- | --- | --- | --- |
|  | N child Pgp3– | Person-years at risk | Incident conversion | SCR | N child Pgp3+ | Person-years at risk | Incident conversion | SRR |
| **Overall** | 1542 | 1432.5 | 219 | 15.3 | 886 | 875.0 | 22 | 2.5 |
| **Study Period** |  |  |  |  |  |  |  |  |
| 0–12m | 566 | 522.5 | 87 | 16.7 | 256 | 251.5 | 9 | 3.6 |
| 12–24m | 536 | 501.5 | 69 | 13.8 | 308 | 303.0 | 10 | 3.3 |
| 24–36m | 440 | 408.5 | 63 | 15.4 | 322 | 320.5 | 3 | 0.9 |
| **Child age** |  |  |  |  |  |  |  |  |
| 0 – 5 y | 1310 | 1214.0 | 192 | 15.8 | 698 | 688.5 | 19 | 2.8 |
| 6 – 9 y | 231 | 217.5 | 27 | 12.4 | 187 | 185.5 | 3 | 1.6 |
| **IgG level†** |  |  |  |  |  |  |  |  |
| [0.0, 3.0] | 1536 | 1427.0 | 218 | 15.3 | 0 | 0.0 | 0 | -- |
| (3.0, 3.5] | 6 | 5.5 | 1 | 18.2 | 47 | 40.0 | 14 | 35.0 |
| (3.5, 4.0] | 0 | 0.0 | 0 | -- | 92 | 89.5 | 5 | 5.6 |
| (4.0, 5.0] | 0 | 0.0 | 0 | -- | 747 | 745.5 | 3 | 0.4 |

\* SCR: Seroconversion rate per 100 person-years, SRR: Seroreversion rate per 100 person-years
† Pgp3 IgG level at the start of the one-year period, in log<sub>10</sub> units of Median Florescence Intensity minus background (MFI-bg) measured on the Luminex platform.

